# Safety and Tolerability of Low-Dose Full-Spectrum Cannabidiol in Long-Term Virally Suppressed Adults with HIV: A Randomized Double-Blind Placebo-Controlled Trial

**DOI:** 10.64898/2026.03.21.26348972

**Authors:** Clémence Couton, Mathilde Wanneveich, Barbara De Dieuleveult, Chloé Robin, Kossi Ayena, Alicia Harry, Hélène Klein, Véronique Avettand-Fenoel, Laurent Hocqueloux, Lucile Mollet, Thierry Prazuck

## Abstract

**Introduction:** People with long-term virologically suppressed HIV (PWH) experience chronic inflammation. Beneficial effects such as lower levels of inflammation were reported for cannabis-based medicine but data on the safety of standardized low-dose full-spectrum cannabidiol (CBD) are limited.

**Methods:** This double-blind randomized placebo-controlled trial (NCT05306249) included 80 ART-treated PWH with undetectable viremia (median time on efficient ART 14 years, median age 54 years), encompassing 30% women. Participants received 1 mg/kg CBD oil twice daily (full-spectrum, THC <0.3%) or placebo for 12 weeks plus a 4-week follow-up. Primary trial endpoint (autophagy gene expression) will be described elsewhere; here we evaluate the treatment impact on prespecified safety outcomes such as hemodynamic with electrocardiograms, HIV immunovirological parameters and comprehensive assessments of liver and kidney functions, performed using standard blood tests. Mixed-effects models adjusted for baseline age, sex, BMI, CD4 count and duration of viral suppression assessed longitudinal changes.

**Results:** Of 80 randomized participants, 35 PWH in CBD and 37 in placebo groups completed week 12. No clinically meaningful differences emerged in creatinine, aminotransferases (ALT, AST), or conjugated bilirubin. Total bilirubin decreased in the CBD arm vs placebo (mixed effect model considering time, group and time*group, adjusted for covariates p=0.046). In exploratory sex-stratified analysis, a significant difference starting at week 12 (−8.0 bpm [95% CI -15.6; -0.4], p=0.0425) and persisting at week 16 (−7.9 bpm [95% CI -14.6; -1.3], p=0.0191) evidences a lower heart rate in men belonging to the CBD group compared to the placebo group; no change in females. There was no change in plasma viral load, cell-associated HIV-DNA levels and CD4/CD8 ratio.

**Discussion:** Low-dose full-spectrum GMP-certified CBD was well tolerated over 12 weeks in virally suppressed people with HIV. Observed reductions in total bilirubin and male heart rate are exploratory and warrant confirmation in adequately powered trials incorporating inflammatory biomarkers and pharmacokinetics.

## Introduction

Without treatment, human immunodeficiency virus (HIV) infection is characterized by sustained viral replication and progressive loss of CD4+ T lymphocytes, leading to AIDS^1^. Nowadays, thanks to highly active antiretroviral therapies (cART), most people living with HIV (PWH) can achieve a plasma viral load below the level of detection within 12 weeks of treatment^2^ and thereby reach a significantly improved life expectancy, nearly matching that of the general population. However, cART does not fully normalize health. Most PWH start cART during chronic infection and residual viral replication persists in some infected cells, hidden in HIV reservoirs: HIV infection becomes a chronic disease. As a consequence, PWH display chronic inflammation and immunosenescence, leading to the onset of early comorbidities associated with age, such as cardiovascular disorders, kidney diseases and certain cancers^3^. HIV-related comorbidities appear to be related to chronic inflammation and PWH worldwide are seeking strategies to promote healthy aging and mitigate the consequences of inflammaging^4^.

Accompanying the two major acidic forms of phytocannabinoids (pCBs), cannabidiol (CBD) and Δ9-tetrahydrocannabinol (THC), more than 120 other molecules have been identified^5^ and are thought to contribute to the entourage effect^6,7^, whereby the biological activity of cannabis extracts is linked to synergistic interactions. This may render full- or broad-spectrum CBD extracts more effective than CBD alone. *In vivo*, pCBs interfere with the endocannabinoid system (ECS)^8^, which is involved in the regulation of numerous physiological and pathological pathways^9,10^, including those of the immune system^11,12^. pCBs act as ligands to the two ECS main receptors: CB1, mainly expressed on cells of the nervous system, and CB2, primarily expressed on immune cells. However, ECS has become more complex, as many other receptors have since been described^9^. Ultimately, pCB receptors are expressed in almost all organs^11^.

Historically, cannabis has attracted significant interest to relieve a range of symptoms in different medical fields, such as cancer and HIV infection^13–15^. Unlike THC, CBD has no psychoactive effect and can be used as a dietary supplement marketed for various potential benefits. In humans, most available studies have focused on recreational cannabis users and have reported effects on inflammation^16–18^. However, in such settings the CBD/THC ratio and the actual doses of absorbed cannabinoids are neither constant nor controlled. A CBD/THC combination (Sativex*®* / Nabiximols) is currently licensed, but Epidiolex*®* is currently the only FDA-approved drug with highly purified CBD as the main active ingredient. This pharmaceutical-grade medicine is prescribed to treat seizures associated with Lennox-Gastaut and Dravet syndromes or tuberous sclerosis complex. In both contexts, when added to conventional antiepileptic regimens, CBD resulted in reductions in the frequency of drop seizures^19,20^. However, among adverse effects, 9% of patients had elevated aminotransferase concentrations, but none met criteria for drug-induced liver injury^20,21^.

In addition, cannabis is known to possess anti-inflammatory properties^22^ and robust clinical trials are needed to evaluate the safety and the ability of cannabis-based medicines (CBM) to alleviate HIV-associated inflammation^23^. Despite encouraging findings in a randomized, open-label, interventional pilot study (CTNPT 028)^24,25^, evidence on the tolerability of CBM in the context of HIV infection remains scarce.

While the primary outcome of the present double-blind randomized, placebo-controlled trial was to assess the effect of a full-spectrum CBD oil (CBD50 LGP CLASSIC, see below) versus placebo on the expression of major autophagy-related genes (manuscript in preparation), we evaluated here the clinical and biological safety and tolerability of oral medical CBD in 80 enrolled PWH as a secondary objective. As part of the French national CBM safety evaluation, we monitored classical laboratory assessments of renal, hepatic and cardiac tolerance of a medical full-spectrum CBD oil at 1 mg/kg, twice daily for 12 weeks.

## Materials and Methods

### Participants

This double-blind, randomized, placebo-controlled trial was conducted at the Department of infectious diseases of the University Hospital of Orléans (France). In addition to the previously reported^26^ inclusion and exclusion criteria, participants were required to have an effectively suppressed (<50 copies/ml) plasmatic viral load for at least 3 years, and no disease or history of severe cardiovascular or cerebrovascular disease, renal failure, severe hepatic impairment, unstable liver disease, cirrhosis or known biliary abnormality.

At W0 (CBD/placebo initiation), urine and blood samples were collected to screen for prior THC and CBD exposure respectively, and 80 adults were enrolled. This study was designed and implemented in accordance with the Declaration of Helsinki (2013 revision) and all patients provided written, informed consent to participate. The protocol was approved by the French Ethics Committee Ouest 6 (#CPP 1431 ME1, 2021-EudraCT 2020-005851) and is registered with Clinicaltrials.gov as NCT05306249. Enrollment began in 05/2022, and the last samples were collected in 02/2023. While the primary outcome of the present trial was to assess the effect of a full-spectrum CBD oil versus placebo on the expression of major autophagy-related genes, we evaluated the clinical and biological safety and tolerability of medical CBD in PWH as a secondary objective. Sample size calculations for the primary endpoint assumed an alpha risk of 5%, a beta risk of 10%, and the standard deviation was calculated from a previous study^27^.

### Trial design and study treatment

The CBM consisted of an orally-administered full-spectrum CBD-dominant extract (THC<0.3%) formulated in medium-chain triglyceride (MCT) oil (CBD 50 mg/mL; CBD50 LGP CLASSIC; Little Green Pharma, Perth, Australia). The placebo was an orally administered formulation consisting of MCT oil. Both products appeared as a thick liquid in a dark 50-mL glass bottle.

Participants were randomly assigned, with a 1:1 allocation ratio, to either receive a double-blind 12-week course of the CBD formulation (1 mg/kg, twice daily) (CBD group hereafter) or the placebo oil, plus a 4-week follow-up. They came to their recruitment center at 4-week intervals (W0, W4, W8, W12, W16). The duration of the washout period was chosen to ensure the clearance of CBD from participants’ plasma^28^. The flowchart of the study is presented in Figure 1.

**Figure 1:**
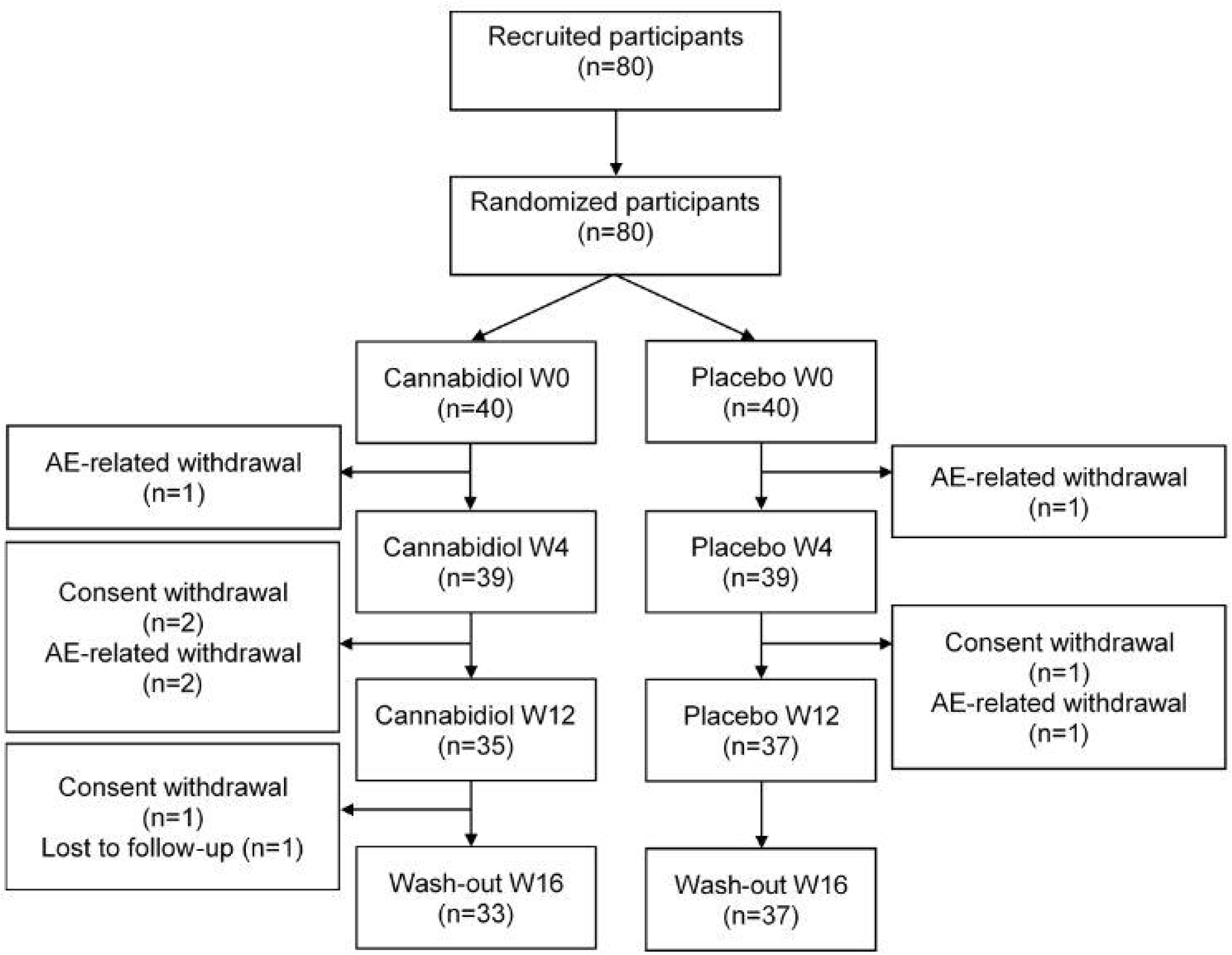
Flow chart of the study. AE, adverse event. W, week. n, number of participants.

### Safety and tolerability assessments

Blood samples were drawn for biological parameters. Complete blood count, CD4 and CD8 T-cells counts and plasma viral load were quantified using standard and automatized procedures in a certified institutional laboratory at W0, W4, W12 (CBD/placebo treatment end) and W16 (follow-up end), aspartate aminotransferase (AST), alanine aminotransferase (ALT) and total and conjugated bilirubin at W0, W4, W8, W12 and creatinine at W0, W4 and W12. Creatinine clearance was calculated using MDRD study equation. HIV-1 DNA was quantified as described previously^29^. Hemodynamic parameters, namely heart rate and electrocardiogram (ECG), were monitored at W0, W4, W12 and W16.

For each participant, we retrieved age, sex at birth, weight and height (for body mass index -BMI-calculation) HIV-related variables such as antiretroviral treatments, CD4 nadir, date of HIV diagnosis and date of the first persisting undetectable viral load (defined as <50 copies/mL). Adverse events and their severity were recorded throughout the study. Quality of life was analyzed in another publication^26^.

### Statistical analyses

Characteristics of the study population at W0 were described and compared between the two treatment groups to identify any potentially relevant imbalances that might warrant adjustment in subsequent models. Continuous variables were analyzed using the Student’s t-test or the Wilcoxon-Mann-Whitney test, depending on distribution; categorical variables were compared using Chi-square test or Fisher’s exact test, when appropriate. Within each group, changes in immuno-virologic parameters over time were assessed using the Friedman test.

To analyze the evolution of clinical outcomes over time according to treatment group, mixed-effects models were used. These models included fixed effects for treatment group, time since inclusion, and their interaction, with a random intercept to account for intra-individual variability. Models were then adjusted for baseline age, sex, BMI, CD4 count and time since the first persistent undetectable viral load (as categorized variable). Exploratory subgroup analyses were also performed by sex.

Model fit was systematically assessed. When necessary, outcome variables were log-transformed to improve model adequacy.

All analyses were implemented in RStudio, using the lmerTest package for mixed-effects models.

## Results

### Participants’ demographics and baseline characteristics

As depicted in the trial flowchart (Figure 1), of the 80 recruited participants, 4 withdrew their consent voluntarily, 1 was lost to follow-up and 5 withdrew because of adverse events. Comprehensive analyses of adverse events, showing no differences between groups, have been previously reported^26^.

Demographics and characteristics at baseline (W0, inclusion), presented and compared in Table 1, were well balanced between groups. Overall, although PWH enrolled in the study were mostly male (69%), 12 women (30%) were included in the CBD arm and 13 (32%) in the placebo arm. There was no significant difference between groups regarding age, time since HIV diagnosis, time since first persistent undetectable viral load, treatments, CD4 nadir, CD4 count, CD4/CD8 ratio, plasma viral load or HIV-1 DNA. Participants were on median 54 years old, had been diagnosed with HIV 19 years earlier and had maintained an undetectable plasma viral load for 14 years.

**Table 1:**
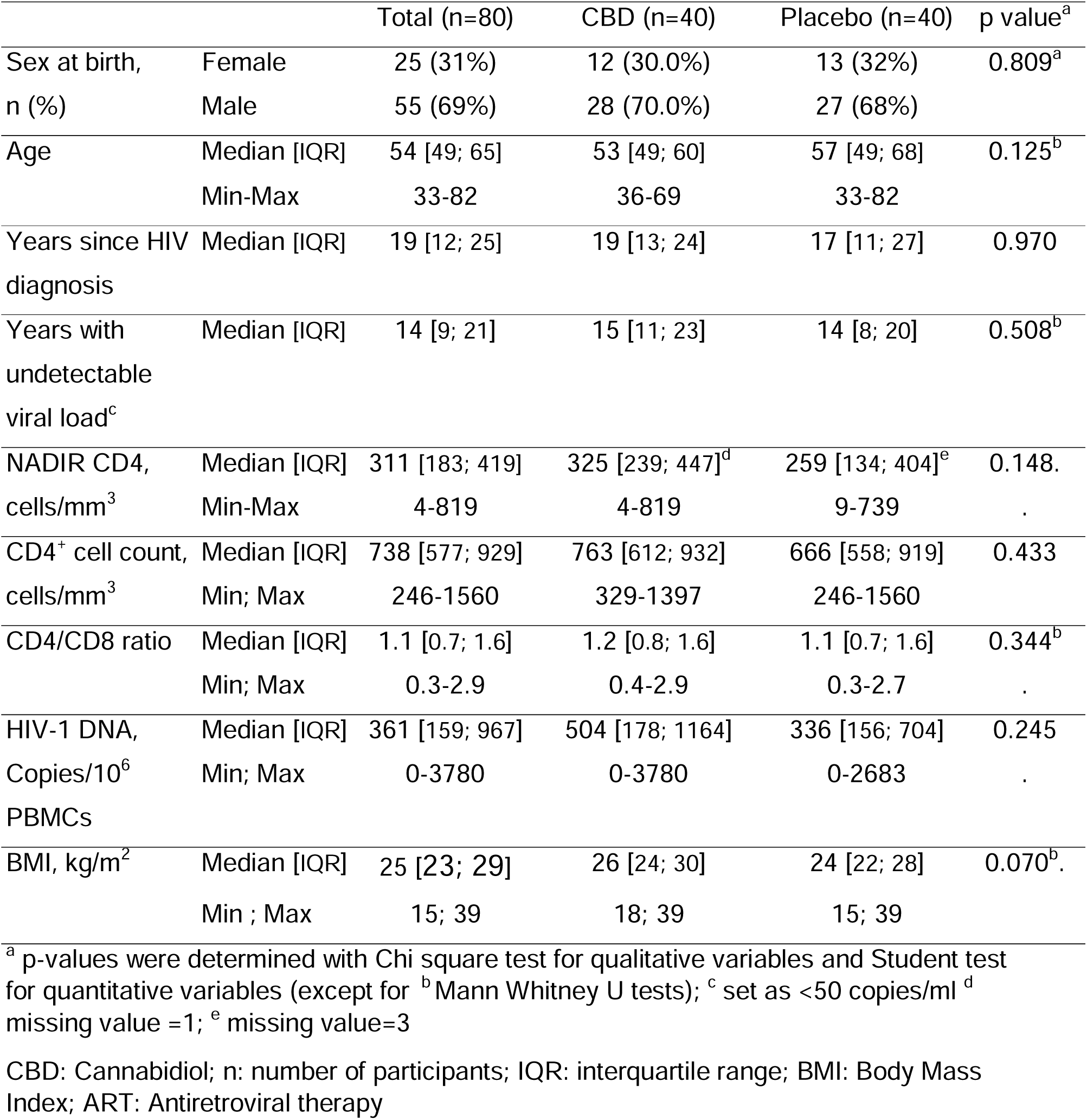
Characteristics of study participants at baseline.

|  |  | Total (n=80) | CBD (n=40) | Placebo (n=40) | p value<sup>a</sup> |
| --- | --- | --- | --- | --- | --- |
| Sex at birth,<br>n (%) | Female | 25 (31%) | 12 (30.0%) | 13 (32%) | 0.809<sup>a</sup> |
|  | Male | 55 (69%) | 28 (70.0%) | 27 (68%) |  |
| Age | Median [IQR] | 54 [49; 65] | 53 [49; 60] | 57 [49; 68] | 0.125<sup>b</sup> |
|  | Min-Max | 33-82 | 36-69 | 33-82 |  |
| Years since HIV<br>diagnosis | Median [IQR] | 19 [12; 25] | 19 [13; 24] | 17 [11; 27] | 0.970 |
| Years with<br>undetectable<br>viral load<sup>c</sup> | Median [IQR] | 14 [9; 21] | 15 [11; 23] | 14 [8; 20] | 0.508<sup>b</sup> |
| NADIR CD4,<br>cells/mm<sup>3</sup> | Median [IQR] | 311 [183; 419] | 325 [239; 447]<sup>d</sup> | 259 [134; 404]<sup>e</sup> | 0.148. |
|  | Min-Max | 4-819 | 4-819 | 9-739 |  |
| CD4<sup>+</sup> cell count,<br>cells/mm<sup>3</sup> | Median [IQR] | 738 [577; 929] | 763 [612; 932] | 666 [558; 919] | 0.433 |
|  | Min; Max | 246-1560 | 329-1397 | 246-1560 |  |
| CD4/CD8 ratio | Median [IQR] | 1.1 [0.7; 1.6] | 1.2 [0.8; 1.6] | 1.1 [0.7; 1.6] | 0.344<sup>b</sup> |
|  | Min; Max | 0.3-2.9 | 0.4-2.9 | 0.3-2.7 |  |
| HIV-1 DNA,<br>Copies/10<sup>6</sup><br>PBMCs | Median [IQR] | 361 [159; 967] | 504 [178; 1164] | 336 [156; 704] | 0.245 |
|  | Min; Max | 0-3780 | 0-3780 | 0-2683 |  |
| BMI, kg/m<sup>2</sup> | Median [IQR] | 25 [23; 29] | 26 [24; 30] | 24 [22; 28] | 0.070<sup>b</sup>. |
|  | Min ; Max | 15; 39 | 18; 39 | 15; 39 |  |

<sup>a</sup> p-values were determined with Chi square test for qualitative variables and Student test for quantitative variables (except for <sup>b</sup> Mann Whitney U tests); <sup>c</sup> set as <50 copies/ml <sup>d</sup> missing value =1; <sup>e</sup> missing value=3
CBD: Cannabidiol; n: number of participants; IQR: interquartile range; BMI: Body Mass Index; ART: Antiretroviral therapy

The median CD4+ T cell nadir was 311 cells/mm^3^ which had increased to 738 cells/mm³ at inclusion in this trial, with a median CD4/CD8 ratio of 1.1, indicating good immune reconstitution. BMI did not differ significantly between groups (p=0.070).

### Immunovirologic parameters

CD4+ and CD8+ T lymphocyte counts (data not shown) and the CD4/CD8 ratio (Figure 2-A) remained stable during the trial. Friedman pairwise comparison tests indicated that none of these parameters changed significantly over the study period, in either group.

**Figure 2:**
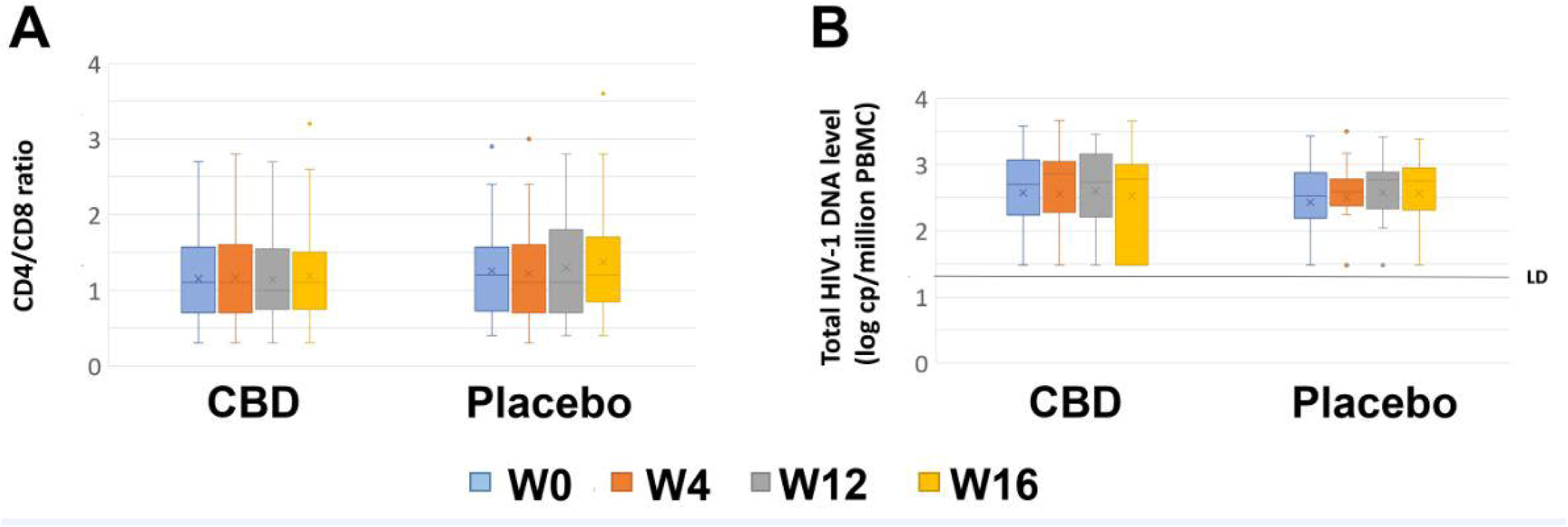
Main immuno-virologic parameters remained stable in both groups. All biologic parameters were obtained using standard procedures in a certified institutional biochemical laboratory and were evaluated at W0 (inclusion, blue), W4 (orange), W12 (end of treatment, grey) and W16 (end of follow-up, yellow). (A) CD4/CD8 ratio was calculated using plasmatic counts of CD4+ T lymphocytes and CD8+ T lymphocytes. (B) Total HIV-1 DNA was quantified by real-time qPCR with a limit of detection (LD) of 30 copies of HIV-1 DNA per million PBMC.

Throughout the study, participants continued antiretroviral treatment, and plasma viral load remained undetectable in both arms (data not shown). The viral reservoir, estimated by HIV-1 DNA level, did not change in either group (Figure 2B).

In summary, HIV-1 proviral DNA levels, undetectable plasma viral load and CD4/CD8 ratio remained stable in all participants, a key issue for PWH.

### Renal tolerance

At baseline, no participants had personal history of renal failure. Kidney function was assessed at W0, W4 and W12 (treatment end). Serum creatinine levels did not change over time or between treatment groups (data not shown). The unadjusted and adjusted mixed-effect models did not reveal any statistically significant effects of treatment group, time or their interaction on clearance (conditional R² around 0.90). Therefore we can conclude that administration of full-spectrum CBD oil at 1 mg/kg twice daily did not alter renal function.

### Hepatic tolerance

Total and conjugated bilirubin and liver transaminases ALT and AST were measured at W0, W4, W8 and W12 and values remained stable overall.

Women had lower levels of ALT (p=0.008), total bilirubin (p=0.001) and conjugated bilirubin (p=0.013) compared to men whereas AST did not differ by sex (p=0.82). However, in males, total bilirubin was significantly reduced after 12 weeks of CBD oil compared with placebo (p=0.0042) (Figure 3 A to C).

**Figure 3:**
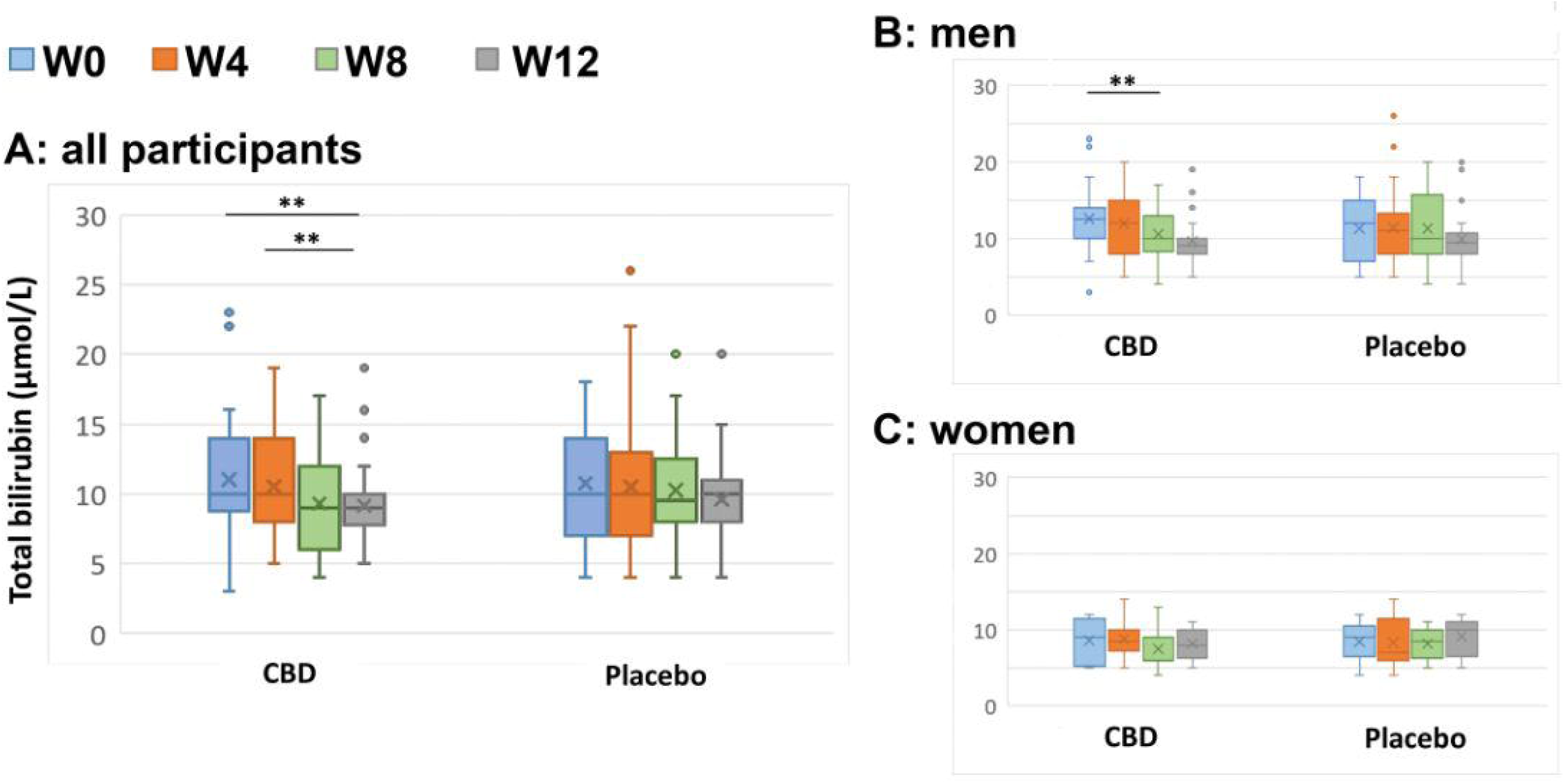
For male participants, full-spectrum CBD managed to significantly decrease plasmatic total bilirubin. Plasma analyses for total bilirubin were compared using Friedman tests considering * p-value< 0.05, ** p-value< 0.01. Data for total bilirubin are represented as (A, left) time*group, and (right) time*group by sex (B: men and C: women)

Using unadjusted and adjusted mixed-effects models, no significant effects of treatment group, time or their interaction were observed on conjugated bilirubin, AST or ALT levels (data not shown), while total bilirubin (log-transformed) was significantly associated with time (p<0.001), showing a decrease over the study period (Conditional R²=0.56). After adjustment for covariates, the effect of time persisted (p=0.001), and both sex and interaction between time and treatment group were associated with changes in total bilirubin levels (p=0.001 and p=0.046, respectively; conditional R²=0.60). More specifically, total bilirubin levels (log-transformed) were higher in males compared to females, and the decline over time was more pronounced in the CBD group than in the placebo group (Table 2). Exploratory subgroup analyses by sex suggested a different temporal trend between treatment groups, that didn’t reach statistical significance.

**Table 2.**
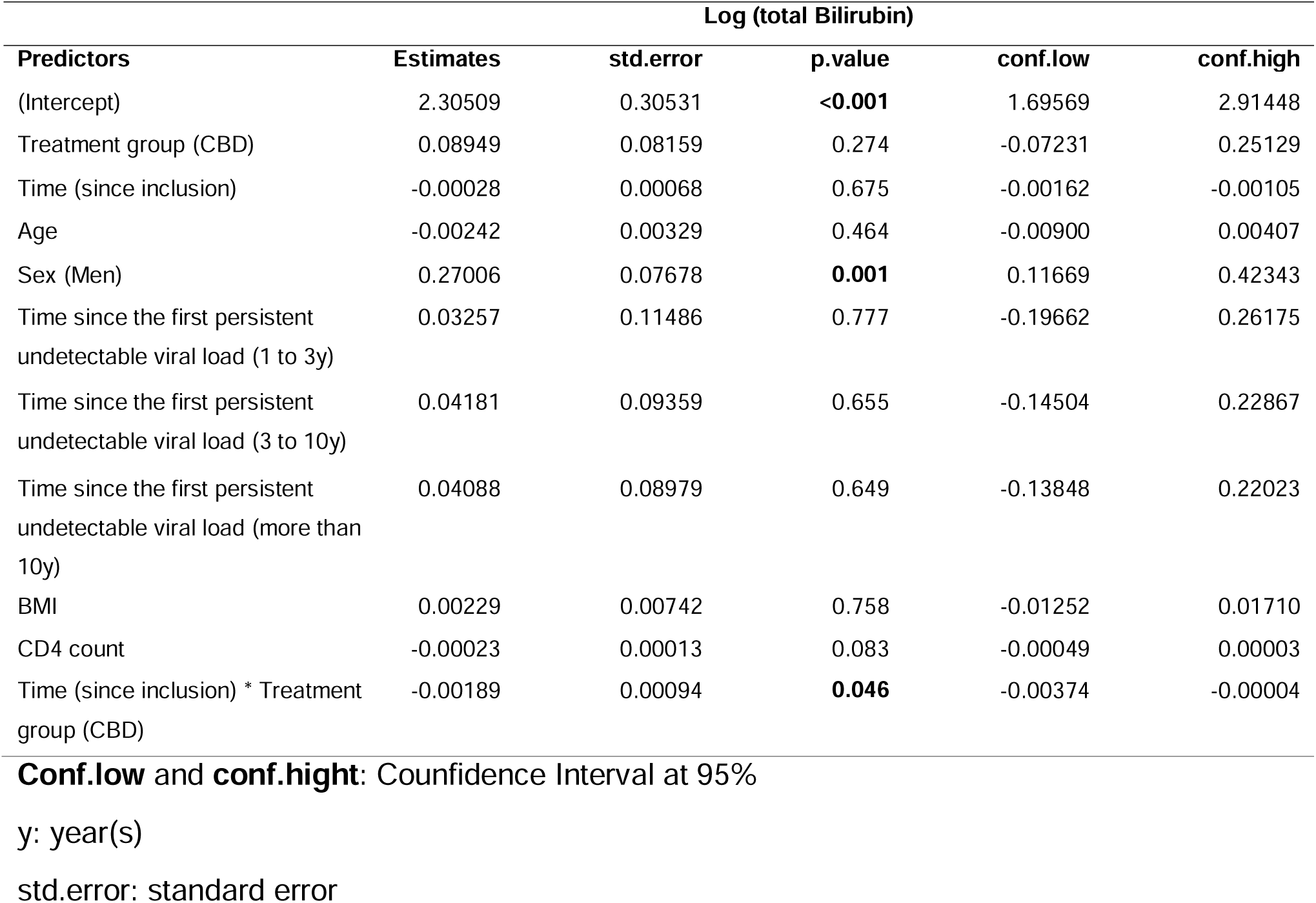
Evolution of total bilirubin over time according to treatment group, adjusted for potential risk factors at baseline.

### Cardiac tolerance

No differences between groups were observed in hemodynamic parameters. Electrocardiograms were considered normal and the few abnormalities detected in both arms were not clinically significant.

Using unadjusted and adjusted mixed-effects models (data not shown), no significant effects of treatment group, time or their interaction were observed on diastolic blood pressure levels. For systolic blood pressure (SBP), a time effect was detected (p=0.012, conditional R²=0.59), indicating an increase over the study period. After adjustment for covariates, the effect of time remained (p=0.015), and both sex and BMI were also associated with SBP levels (p=0.031 and p=0.002 respectively, conditional R²=0.61). More specifically, SBP values were higher in men than in women and increased with higher BMI. In exploratory subgroup analyses by sex, no association was found between time, treatment group or their interaction on SBP in women. However, in men, both time and BMI were associated with higher SBP levels (p=0.006 and p=0.02 respectively, conditional R²=0.61).

Regarding heart rate, the CBD group had a significantly lower heart rate than the placebo group at week 12 (67 bpm vs. 73 bpm respectively; p=0.035), and this difference persisted four weeks later (68 bpm vs. 75 bpm, p=0.027). In exploratory subgroup analyses by sex, this difference was observed only in men (Figure 4): at week 12, median heart rate was 64 bpm in CBD group vs 73 bpm in placebo group (p=0.043), and at week 16, 63 bpm vs 78 bpm, respectively (p=0.019). These results suggest a reduction in heart rate over time among men in the CBD group.

**Figure 4:**
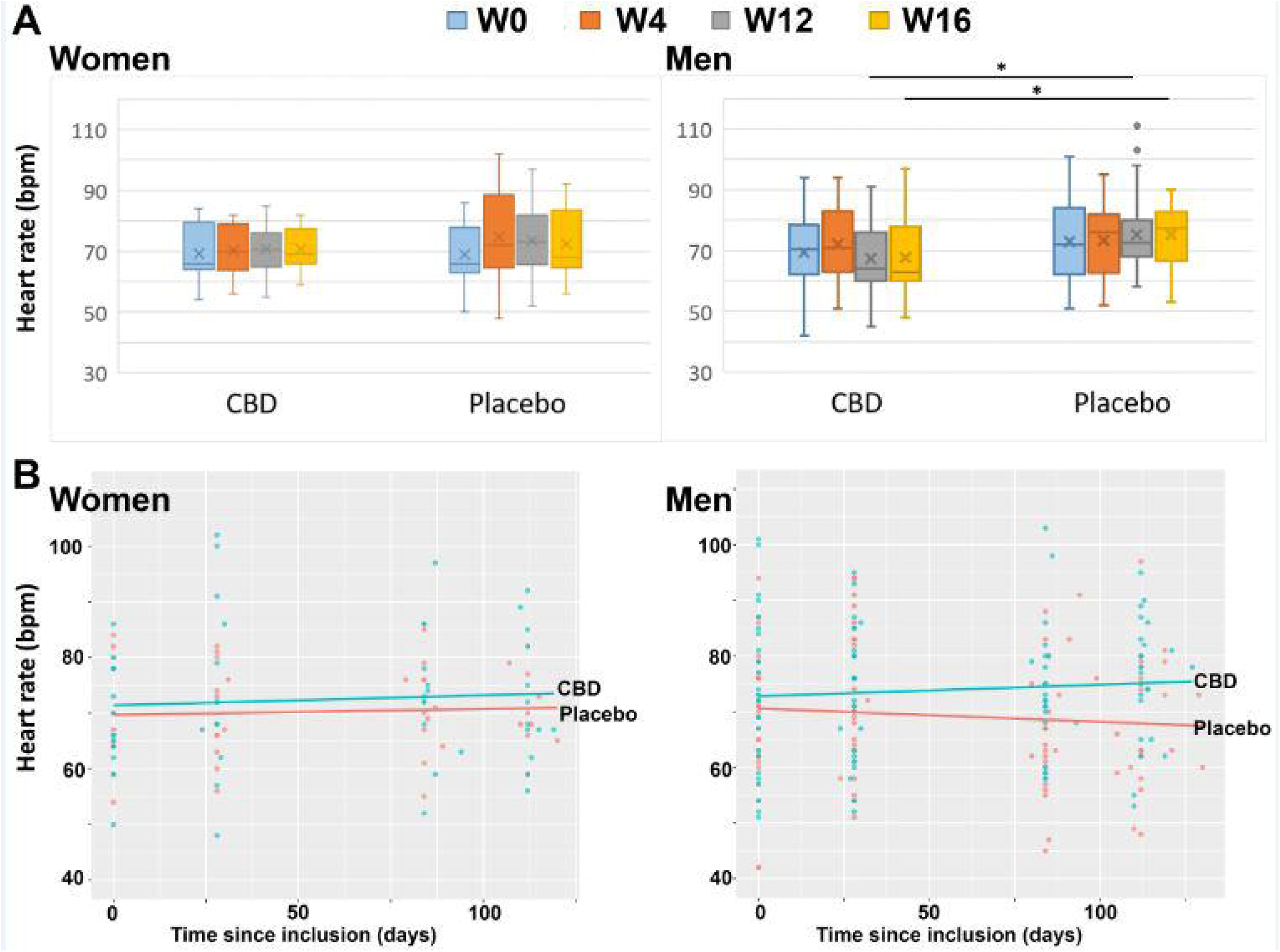
full-spectrum CBD significantly decreased heart rate in men but not in women living with HIV. Heart rate (bpm, beats per minute) was measured at W0 (inclusion), W4, W12 (end of treatment), and W16 (4 weeks post-treatment/washout) and comparison was made between female (left) and male (right) in both groups (placebo and CBD). A: Group comparisons for each time point were conducted using an unpaired Student’s t-test, with p<0.05 (*) defining statistical significance. B: Time-course of heart rate change following CBD or placebo administration, stratified by sex. The left panel displays data for females; the right panel displays data for males. Orange circles/lines indicate CBD treatment; blue circles/lines indicate placebo treatment.

## Discussion

In this double-blind, randomized, placebo-controlled trial of adults with long-term virologically suppressed HIV, a stable, high-quality GMP-certified, full-spectrum cannabidiol (CBD; 1 mg/kg twice daily; THC < 0.3%) adjusted to each participant’s body weight, was well tolerated over 12 weeks, with no clinically meaningful between-group differences in renal, hepatic, or cardiac laboratory parameters, and no sign of immunovirologic harm. These findings were consistent across adjusted mixed-effects analyses and aligned with the study’s prespecified safety endpoints. Together with the absence of excess adverse events reported separately^26^, our results support the short-term tolerability of standardized low-dose CBD in PWH. They extend a sparse but growing clinical literature on cannabinoids in HIV, which until recently relied largely on observational cohorts or small pilot trials with heterogeneous products and dosing^23–25,30,31^.

To the best of our knowledge, this is the first randomized, placebo-controlled trial assessing the safety of a pharmaceutical-grade, full-spectrum CBD oil in 80 long-term suppressed PWH. Published in 2022, the only prior trial available is a Canadian open-label clinical study^24,25^ which was designed to include 26 PWH, but only 10 individuals were ultimately enrolled and participants, randomized into two groups (n=5 per group), the first group received THC:CBD capsules (2.5:2.5-15:15 mg/day), while the second group received CBD alone (200-800 mg/day).

Our trial differs from prior work by administering a weight-based, pharmaceutical-grade, full-spectrum CBD with rigorous blinding, placebo control, and serial objective safety measurements (renal, hepatic, cardiac). The use of a standardized product mitigates variability inherent to consumer CBD preparations and strengthens inference on tolerability in PWH^24,25,30,31^. Notably, even if the sample size was not calculated to compare the impact of CBD between men and women, 30% of participants were women, an uncommon proportion in HIV clinical and translational studies where women remain consistently underrepresented. Such repartition permitted us to collect data concerning CBD safety in women as in men. Biological sex is now recognized as an important variable to consider^32^, with numerous differences reported between men and women in HIV pathogenesis, treatment response and non-AIDS comorbidities such as cardiovascular disease^33^. Ensuring adequate female enrolment is therefore critical to improving the evidence base and optimizing care for women with HIV.

Hepatic safety warrants specific comment. Elevations in aminotransferases have been described primarily with high-dose purified CBD (e.g., 10–20 mg/kg/day or more in epilepsy programs) and are uncommon at lower doses in non-HIV settings^20,21,30,31,34–37^. In our trial, aminotransferases remained stable, and conjugated bilirubin did not differ from placebo. We observed, however, a statistically significant reduction in total bilirubin in the CBD arm at week 12 versus placebo. This signal should be considered exploratory: the study was not powered for biochemical efficacy outcomes, and we did not include pharmacokinetic measures or mechanistic biomarkers to probe potential pathways. Replication in longer trials with predefined hepatic endpoints and CBD exposure measurements will be needed to determine whether this observation is reproducible and clinically relevant^30,31,37,38^.

Cardiovascular assessments showed overall stability of blood pressure and ECG parameters. In sex-stratified analyses, men assigned to CBD exhibited a reduction in resting heart rate at W12 that persisted 4 weeks after discontinuation, whereas no change was seen among women. Given the modest sample size, post-hoc nature of this analysis, and known sex differences in autonomic responses to cannabinoids, this should be viewed as hypothesis-generating^39^. Future adequately powered trials that integrate autonomic measures (e.g., heart-rate variability) and inflammatory biomarkers are warranted, particularly in light of the elevated cardiovascular risk in PWH—including women, who remain understudied^33,40^.

Strengths of this trial include the randomized, placebo-controlled design; use of a low-dose, full-spectrum, pharmaceutical-grade CBD oil formulation; serial objective laboratory and hemodynamic evaluations; and inclusion of both men and women with long duration of HIV control on ART (median 15 years). HIV-related parameters (HIV-1 RNA, total HIV-1 DNA, CD4/CD8 ratio) remained stable, which is reassuring against immunovirologic harm over 12 weeks and aligns with recent work using standardized cannabinoid products in PWH^23,25^.

Several limitations temper interpretation. First, the dose was intentionally low and fixed (1 mg/kg twice daily) without up-titration^41^. This gradual titration can influence both tolerability and detection of effects^41^. Second, the 12-week treatment and 4-week washout may be insufficient to reveal uncommon or delayed adverse effects. Third, the study was not powered for subgroup or sex-specific outcomes; the bilirubin and male heart-rate findings require confirmation with prespecified hypotheses and appropriate multiplicity control. Fourth, pharmacokinetic data for CBD and comprehensive inflammatory panels were not part of this safety analysis; integrating these measurements will be critical to link exposure, biological activity, and safety^30,31,41^. Finally, generalizability is limited to PWH with long-term viral suppression and without advanced hepatic or renal disease at inclusion.

## Conclusion

In summary, standardized low-dose full-spectrum CBD was well tolerated over 12 weeks in adults with long-term suppressed HIV, with no evidence of renal, hepatic, cardiac, or immuno-virologic harm. The observed reductions in total bilirubin and in resting heart rate among men are exploratory and require confirmation in larger, longer trials designed to assess mechanistic pathways (inflammation, autonomic tone), pharmacokinetics, and potential drug–drug interactions with antiretrovirals^23–26,30,31,37,39–41^. Pending such data, prudent liver-function monitoring remains advisable when prescribing CBD, especially at higher doses or in patients with pre-existing hepatic comorbidity^20,21,30,31,34–36,41^.

## Data Availability

All data produced in the present work are contained in the manuscript

## Acknowledgments,

We thank all study participants and hospital staff who participated in the trial.

## Authorship confirmation/contribution statement (CRediT format is preferred)

Clémence Couton: Formal analysis, Investigation, Writing - original draft, Writing – review and editing, Visualization.

Mathilde Wanneveich: Formal analysis, Writing – review and editing, Visualization.

Barbara De Dieuleveult: Investigation, Writing – review and editing.

Chloé Robin: Investigation, Writing – review and editing. Hélène Klein: Resources.

Kossi Ayena: Investigation, Writing – review and editing.

Véronique Avettand-Fènoël: Resources, Writing – review and editing.

Laurent Hocqueloux: Resources, Writing – review and editing.

Lucile Mollet: Conceptualization, Writing - original draft, Writing – review and editing, Supervision, Funding acquisition.

Thierry Prazuck: Conceptualization, Resources, Writing – review and editing, Supervision, Funding acquisition.

